# Disentangling craving- and valence-driven brain responses to smoking cues in individuals with nicotine use disorder

**DOI:** 10.1101/2020.08.10.20171611

**Authors:** Amelie Haugg, Andrei Manoliu, Ronald Sladky, Lea M. Hulka, Matthias Kirschner, Annette B. Brühl, Erich Seifritz, Boris B. Quednow, Marcus Herdener, Frank Scharnowski

## Abstract

Tobacco smoking is one of the leading causes of preventable death and disease worldwide. Most smokers want to quit, but relapse rates are high. To improve current smoking cessation treatments, a better understanding of the underlying mechanisms of nicotine dependence and related craving behavior is needed. Studies on cue-driven cigarette craving have been a particularly useful tool for investigating the neural mechanisms of drug craving. Here, functional neuroimaging studies in humans have identified a core network of craving-related brain responses to smoking cues that comprises of amygdala, anterior cingulate cortex, orbitofrontal cortex, posterior cingulate cortex, and ventral striatum.

However, most functional Magnetic Resonance Imaging (fMRI) cue-reactivity studies do not adjust their stimuli for emotional valence, a factor assumed to confound craving-driven brain responses to smoking cues. Here, we investigated the influence of emotional valence on key addiction brain areas by disentangling craving- and valence-related brain responses with parametric modulators in 32 smokers. For one of the suggested key regions for addiction, the amygdala, we observed significantly stronger brain responses to the valence aspect of the presented images than to the craving aspect. Our results emphasize the need for carefully selecting stimulus material for cue-reactivity paradigms, in particular with respect to emotional valence. Further, they can help designing future research on teasing apart the diverse psychological dimensions that comprise nicotine dependence, and, therefore, can lead to a more precise mapping of craving-associated brain areas, an important step towards more tailored smoking cessation treatments.

## Introduction

Tobacco smoking is one of the leading risk factors for preventable death and disease worldwide and it is estimated to kill 8 million users per year (1-3). Even though the number of nicotine dependent smokers who want to quit smoking is large (4), many smokers are not successful with their smoking cessation goals, and relapse rates are high (5-7). In addition, the efficacy of available psychological and pharmacological treatments against nicotine dependence is still limited (8). Causes for smoking relapse are diverse, and, to date, a wide range of factors that have an effect on relapse rates have been identified, such as perceived stress (9), impulsivity (10), or low self-efficacy (11).

Another important factor that influences success in quitting to smoke is the individual’s smoking-related craving (12,13) and the ability to control this craving (14,15). Craving, or, more precisely, drug craving, is commonly described as the “desire to use a drug” (16), even though precise operational definitions can still vary largely across studies (17). In particular, Shiffman and colleagues observed cigarette craving after waking up to be predictive for smoking relapse in smokers who wanted to quit (18). Consequently, encountering social situations associated with smoking and other craving inducing environments pose serious problems for smokers to reach their goal of sustained abstinence. Therefore, cigarette craving has been intensively studied with different methods (16,17). For example, self-report questionnaires, which can vary from a single visual analogue scale (19) to more complex, multi-dimensional questionnaires such as the Questionnaire on Smoking Urges (20) have been established as suitable methods to assess baseline craving levels. Another important approach that focuses more on situationally-induced craving is the so-called cue-reactivity paradigm (21–23). Here, smokers are exposed to smoking cues, such as real cigarettes or visual stimuli that depict smoking scenes, and their subjective, behavioral, or physiological craving response is measured. Cue-reactivity paradigms combined with neuroimaging are particularly suitable for identifying human brain responses associated with craving (24–26).

Recent meta-analyses of cue-reactivity functional Magnetic Resonance Imaging (fMRI) studies found relatively robust drug cue-driven brain activity in the ventral striatum, amygdala, anterior cingulate cortex (ACC), posterior cingulate cortex (PCC), and orbitofrontal cortex (OFC) (24–26) (see Figure 1). These brain areas were repeatedly identified in cue-reactivity studies and may play an important role in the neural mechanisms underlying substance use disorders (27). For instance, the ventral striatum is considered as one of the most prominent reward-associated areas, and has been directly linked to reward processing and reward learning in animal and human studies (28,29), making it a key area involved in addiction. Also, the amygdala, next to its strong involvement in emotional processes (30), also mediates Pavlovian processing, making it a crucial instance in the learning of associations between environmental cues and drug-induced reward and, therefore, in reinforcing drug seeking behavior (31,32). The ACC, PCC, and OFC, known to be involved in higher order cognitive processes, have also been linked to neural mechanisms related to substance use disorder: The ACC as a particularly strongly connected brain area is associated with decision-making as well as executive control, and exerts top-down control over reward-driven areas such as the ventral striatum (33). The OFC is involved in evaluating the value of rewards and, therefore, another important area for reward processing (34). Finally, the PCC has been observed to be more indirectly involved in substance use disorder. The PCC is part of the default mode network and associated with self-related processes, being activated when resisting cue-driven craving (35).

**Figure 1:**
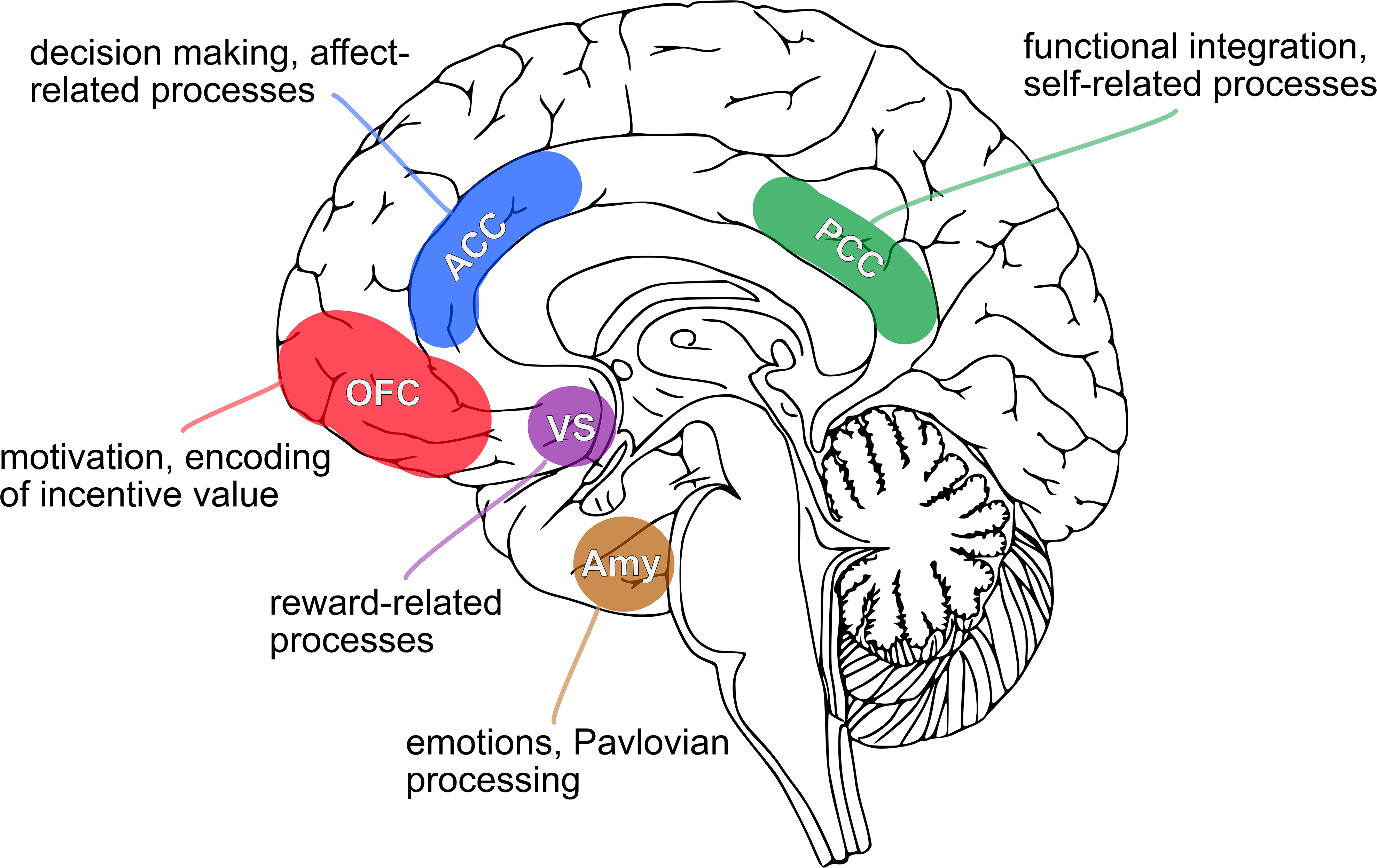
Most commonly observed brain areas in fMRI-based cue-reactivity studies. Cue-reactivity meta-analyses (24–26) identified a wide range of different brain areas driven by drug cues, with the amygdala (Amy), the ventral striatum (VS), the anterior cingulate cortex (ACC), the orbitofrontal cortex (OFC), and the posterior cingulate cortex being the most robustly identified cue-driven brain areas.

Cue-reactive brain responses have also been shown to be predictive of smoking abstinence and relapse (36,37), thus, further underlining the clinical importance of understanding the neural mechanisms in these brain areas regarding nicotine dependence and craving.

The multifaceted neural underpinnings of smoking cue-reactivity reflect the many psychological dimensions that are associated with the perception and processing of such cues. Smoking cues are associated with the urge to smoke as well as factors such as the positive or negative connotation of the depicted situation. Consequently, the unique contribution of smoking-related craving to measured neural responses can be confounded by other stimulus dimensions that, as well, influence addiction-related brain areas. In particular, the emotional dimension of smoking cues can potentially confound craving-associated responses, as affective stimuli have been shown to strongly engage brain areas like the amygdala (38,39), an area that has also been identified in a wide range of cue-reactivity studies (24-26). This is of particular relevance as the majority of fMRI-based cue-reactivity studies (e.g. 80% of the fMRI studies analyzed in the aforementioned three meta-analyses) used a classical fMRI block design to identify brain areas that are driven by smoking-associated cues: blocks of drug-related images are presented in an alternating fashion with blocks of neutral images. An fMRI block design provides considerably large statistical power to detect effects, but its inflexible design of a priori selected images for each stimulus block does not allow for further analyses with respect to other stimulus characteristics (40,41). This inflexibility can, consequently, be problematic when other stimulus characteristics (other than the ones chosen for designing the stimulus blocks) are driving different/additional brain dynamics. For instance, craving-related brain regions might also be driven by other aspects of the presented smoking cues, such as their emotional content.

To overcome this limitation, we applied an event-related parametric fMRI design combined with ratings of craving and valence as behavioral measures to disentangle core components of craving- and valence-driven brain regions. This allowed us to identify key regions within the “addiction network” (27) that were more sensitive to and more driven by either the craving or the emotional content of the presented craving cues. A better understanding of specific craving-as well as valence-driven brain activations can help to optimize future cue-reactivity paradigms and, more importantly, can help to identify more reliable targets for clinical interventions, such as cue-exposure therapy or real-time fMRI neurofeedback.

## Material and Methods

### Participants

32 subjects with nicotine use disorder (age: 25.93±5.30; 16 females, 15 males, 1 non-binary; average daily cigarette consumption of 11.47±5.57 cigarettes; smoking duration of 7.41±4.76 years) participated in the study. All subjects gave informed written consent and were compensated for their participation in the study (25CHF/hour). Exclusion criteria were mental or neurological disorders, MRI-incompatibility criteria (i.e., metal implants, current pregnancy, pacemakers, etc.), as well as the use of any non-cigarette tobacco substitutes, such as nicotine patches, chewing gums, or electronic cigarettes. Inclusion criterium was tobacco use disorder according to DSM-5 (42).

### Experimental procedure and design

All subjects were instructed to abstain from smoking at least one hour before the study. Prior to scanning, subjects were asked to fill out several questionnaires which included a drug use anamnesis of current and past drug use (previously described in Quednow, Kühn, Hoenig, Maier, & Wagner, 2004), the Fagerström Test for Nicotine Dependence (44), and the brief Questionnaire of Smoking Urges (QSU) (45). The QSU was filled out a second time in the end of the experimental session.

In the MRI session, subjects underwent a passive viewing paradigm (code for image presentation available at https://osf.io/6y8fu/) where they were presented a total of 330 neutral and nicotine-related images, distributed over five runs of 5 minutes each (see Figure 2). During each run, 68 images were presented for 2.3 seconds in random order, each followed by a 1-second fixation dot as baseline. In the beginning of each run, a 15-second fixation dot baseline was presented. To ensure that all images were passively viewed by the subjects and subjects were paying attention to the images, 10 additional catch trial images were randomly presented during the five runs. Participants were instructed to perform a button press when a catch trial image, depicting an exclamation mark, was presented. All images were taken from the Smoking Cue Database (SmoCuDa) (46), the International Affective Picture System (IAPS) database (47), as well as the International Smoking Image Series (ISIS) database (48) and covered a continuous range from very mild to very intense craving-inducing content. Before and after the passive viewing runs, participants also performed 7-minute resting state scans where they had to fixate a white fixation dot over a black background. After the functional imaging we acquired an anatomical image. In total, one scanning session took approximately 50 minutes.

**Figure 2:**
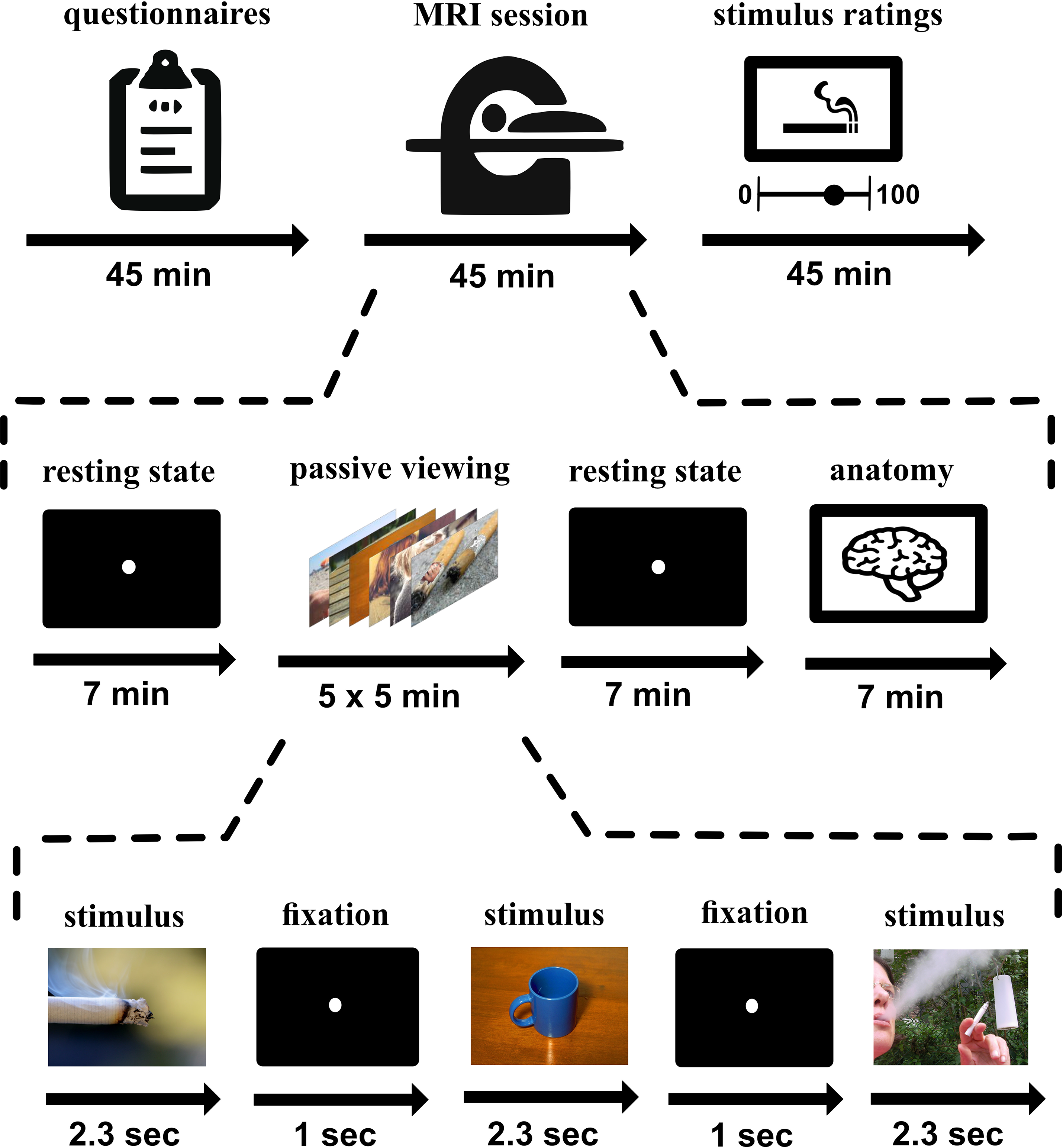
Experimental design. The study was divided into three parts. In the first part, subjects filled out several questionnaires on their smoking routines. Then, subjects underwent 50min of scanning, including resting state scans, 5 runs of passive viewing, and an anatomical scan. During each passive viewing run, 68 out of a total of 340 neutral or nicotine-related images (including 10 additional catch trial images) were presented for 2.3 seconds in random order, followed by a 1-second fixation dot baseline. Finally, subjects rated all 330 presented images with respect to craving and valence.

After scanning, subjects were asked to rate the 330 images that were presented during the passive viewing paradigm on a 100-point visual analogue scale outside the scanner. All images were presented in random order and had to be rated on two dimensions: the subject’s urge to smoke when seeing the image (craving), as well as how positively or negatively the subject perceived the image (valence). A detailed description of the rating procedure can be found in Manoliu et al. (46).

### MRI data acquisition

MR images were acquired with a 3 Tesla Philips Achieva scanner (Philips Healthcare, The Netherlands) using a 32-channel head coil at the MR Center of the Psychiatric University Hospital in Zurich, Switzerland. Functional images during the passive viewing paradigm were acquired using a T2*-weighted gradient-echo planar imaging (EPI) sequence with repetition time (RT) = 2000ms, echo time (TE) = 35 ms, flip angle (FA) = 82°, 27 slices in ascending order, interslice gap = 1mm, voxel size = 2 × 2 × 3 mm^3^, and field of view (FoV) = 220 × 220 × 109 mm^3^. 5 dummy scans as well as 122 functional images were collected during each 4:18min passive viewing run. Further, a T1-weighted sequence in the end of the session was acquired with FA = 8°, 237 sagittal slices in ascending order, no slice gap, voxel size = 0.76 × 0.76 × 0.76, and FoV = 255 × 255 × 180 mm^3^. The anatomical run took 8:26 minutes.

### fMRI analysis

All functional MR images were analyzed using MATLAB2017a and Statistical Parametric Mapping (SPM12; Wellcome Trust Centre for Neuroimaging, London, United Kingdom). Preprocessing of the functional images included: slice-time correction, realignment, co-registration of the functional scans to the anatomical image, segmentation, normalization into Montreal Neurological Institute (MNI) space, and smoothing with a Gaussian kernel of 6mm full width at half maximum.

Before the parametric analysis, we applied a more standard cue-reactivity analysis without parametric modulation to replicate previous approaches. For this, we first specified a general linear model (GLM) with three regressors for high and low craving-intensity smoking stimuli as well as neutral stimuli, and included the six motion parameters as well as the catch trials as regressors of no interest. We then contrasted smoking images that were rated as highly craving inducing, defined as images that received a craving rating of 50 or higher, with neutral images (high-craving-vs-neutral contrast).

For our parametric analyses, we specified two GLMs, GLM-craving and GLM-valence. Each GLM contained the six motion parameters as regressors of no interest and one regressor representing the presented images, modelled as a boxcar function convolved with the hemodynamic response function. GLM-craving additionally contained one parametric modulator for the craving rating for each presented stimulus while GLM-valence additionally contained one parametric modulator for the valence rating for each presented stimulus. In both GLMs, catch trials were modelled as an additional regressor of no interest. Contrast images representing the craving and valence regressor, respectively, were used for second level analyses.

For the region of interest (ROI)-based analysis, we extracted average values from the beta images of the craving and valence parametric regressor of the first level analyses, for each ROI, using custom MATLAB scripts (all used scripts can be found on the Open Science Framework: https://osf.io/6y8fu/). The ROIs were defined based on fMRI cue-reactivity meta studies (24-26) and included the ventral striatum, amygdala, orbitofrontal cortex, anterior cingulate cortex, and posterior cingulate cortex (see supplemental material for a detailed description of ROI selection and definition and the Open Science Framework repository for the mask files: https://osf.io/6y8fu/). One meta-analysis did not differentiate between cue-reactivity studies on different substance use disorders (24), for the other two meta-analyses, we focused on results based on smoking cue-reactivity only. We compared the extracted craving- and valence-beta values using paired t-tests. To investigate brain-behavior relationships, we performed Spearman correlation analyses between extracted beta values and Fagerström dependence scores as well as the number of daily smoked cigarettes, respectively.

### Behavioral analysis

Ratings of the parametric modulators were based on average ratings across 40 nicotine dependent subjects, including the 32 participants of this study, and were performed after the scanning session. A detailed description of the rating procedure and rating distributions has been published in Manoliu et al. (46).

### Statistical analyses

All statistical cluster-level analyses of fMRI data were performed using an initial cluster defining threshold of p<0.001 and family-wise-error (FWE) corrections of 0.05. Paired t-tests between craving- and valence-betas were corrected for multiple comparisons using Bonferroni corrections.

### Code and data availability

The used smoking stimuli, a more detailed overview of stimuli ratings, and the exact scripts used for performing stimulus ratings are all available on the Open Science Framework (OSF) platform (https://osf.io/6gwy5/). Further, we have made all scripts used for fMRI analyses and image presentation publicly available in another repository on the OSF platform (https://osf.io/6y8fu/). This repository also includes group-level neuroimaging data, while individual data were not allowed to be shared publicly by our local ethics regulations.

## Results

### Brain response to high-craving smoking images as compared to neutral images

To replicate conventional cue-reactivity analyses, we contrasted high-craving-vs-neutral, depicting brain areas driven by high-in-craving-rated smoking images as compared to neutral images. Our analysis revealed significant (0.05 FWE-corrected) activation in the higher order visual cortices, the medial prefrontal cortex and ACC, and the PCC (see Figure 3; Table S1 in the Supplemental Material for details).

**Figure 3:**
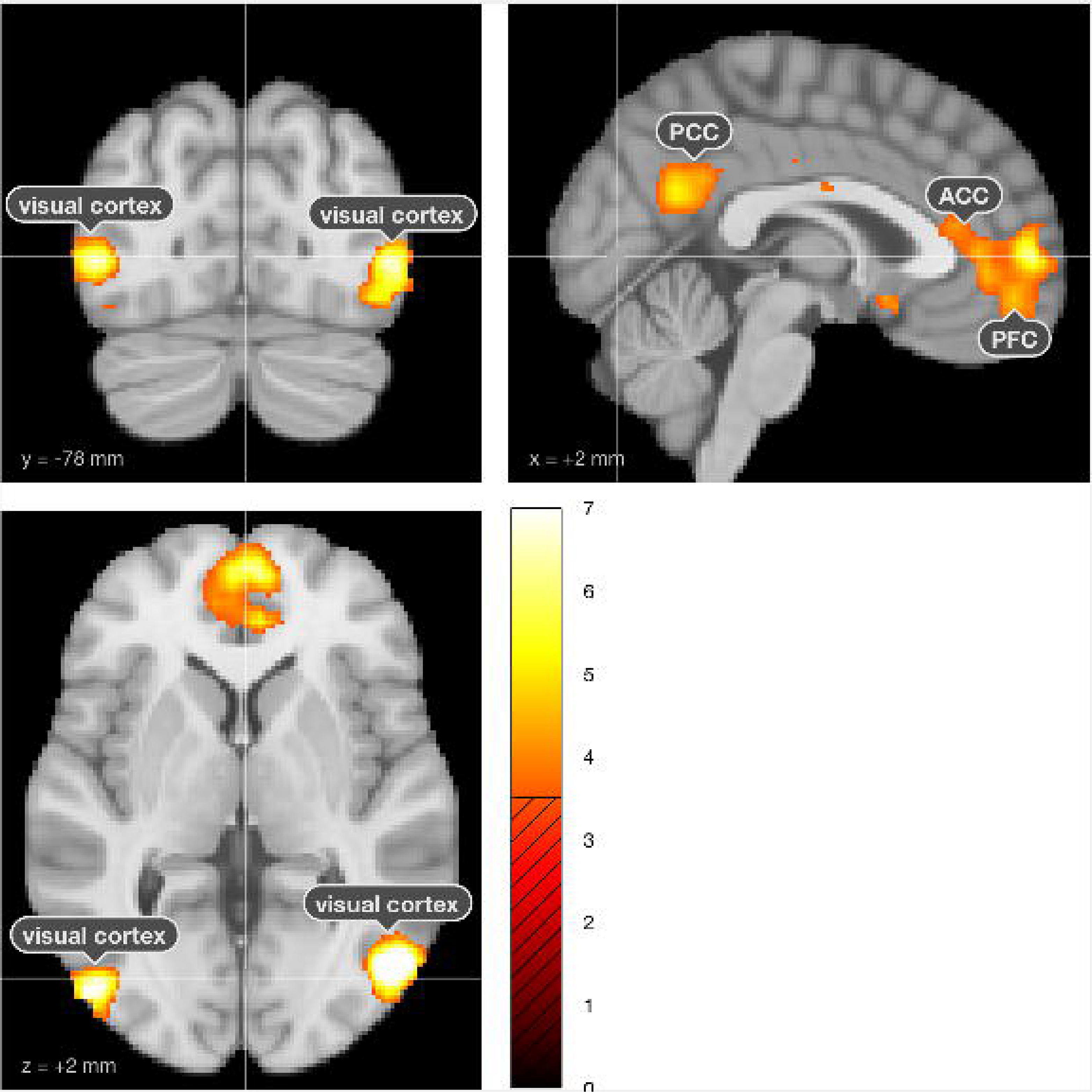
Whole-brain analysis results depicting brain areas activated by high-craving smoking images as contrasted to neutral images. The high-craving-vs-neutral contrast revealed activation in the higher order visual cortices, the prefrontal cortex (PFC) and anterior cingulate cortex (ACC), and the posterior cingulate cortex (PCC).

### Disentangling craving and valence activations

To disentangle brain activations associated specifically with craving or valence, we parametrically analyzed responses to the craving and valence rating, respectively. Whole brain analyses revealed significant craving-related activation within the visual cortex, the PFC and ACC, the PCC, the parietal cortex, the fusiform gyrus, and the precuneus, while significant valence-related activation was observed for the visual cortex and the parietal cortex. More details can be found in the supplemental material (Figure S1, Figure S2 Table S2, Table S3).

In a second step, we specifically investigated craving- and valence-associated activity within our predefined ROIs. The craving regressor showed significant small volume-corrected activation in the OFC, the ACC, the ventral striatum, as well as the PCC (Figure 4; Table S4 in Supplemental Material). In contrast, the valence regressor only showed significant small volume-corrected activation within the amygdala. Statistical comparisons using paired t-tests showed a significant difference between the craving- and valence-driven responses in the amygdala, with valence-driven amygdala activity being significantly higher than craving-driven activity (t(31) = 4.41, p < 0.001; Figure 4). The other four investigated ROIs did not show significant differences between craving- and valence-driven brain responses. Nevertheless, in these four ROIs, craving-driven brain responses were higher than valence-driven brain responses with the latter often reaching values close to zero (see Figure 4). Further details can be found in the supplemental material, Table S4.

**Figure 4:**
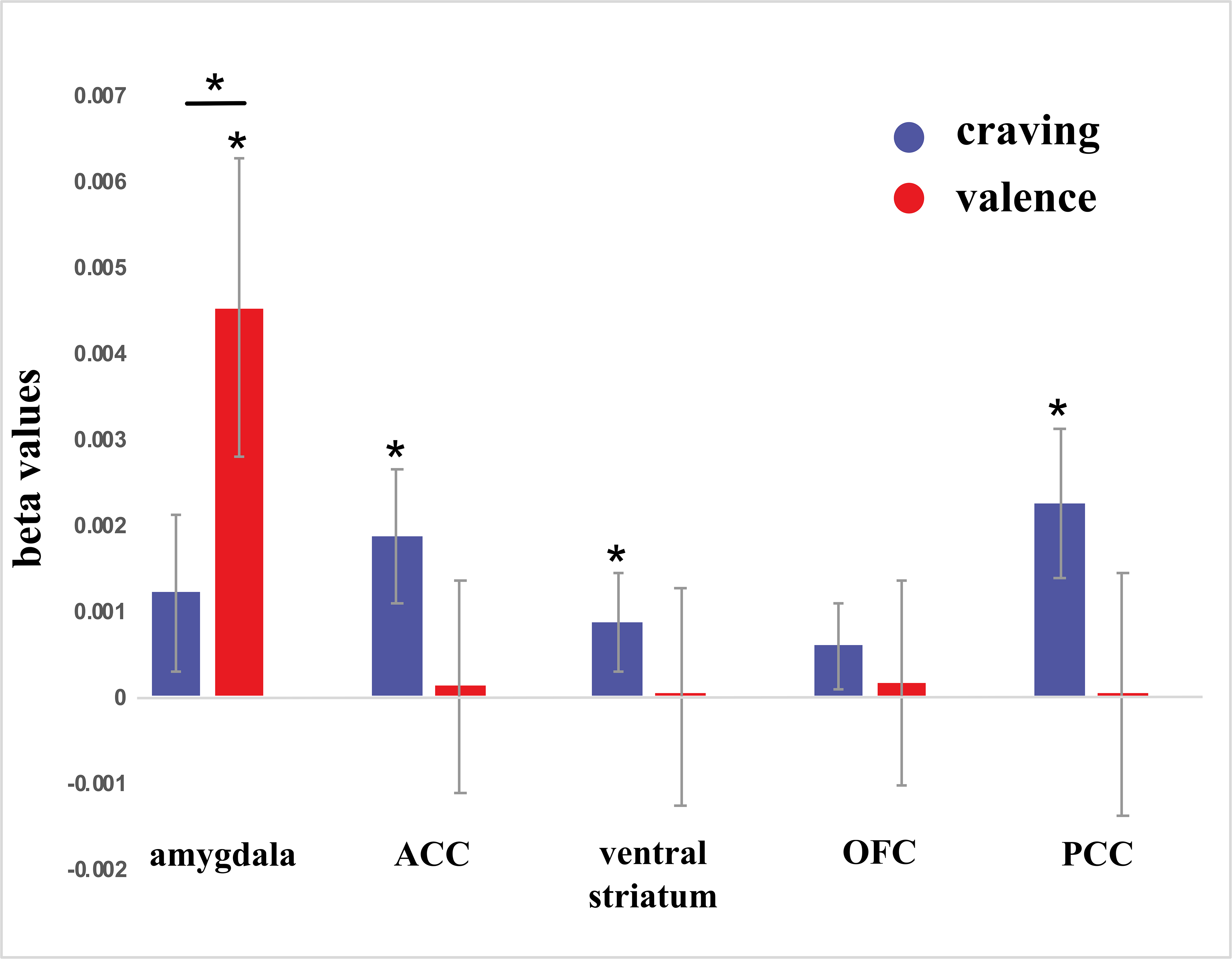
Brain responses to the craving and valence ratings of the presented images. Group-level analyses revealed significant craving-driven activation within the ACC, ventral striatum, and PCC, and significant valence-driven activation within the amygdala. We observed a significant difference between craving- and valence-driven activation within the amygdala. Abbreviations: anterior cingulate cortex (ACC), orbitofrontal cortex (OFC), posterior cingulate cortex (PCC)

## Discussion

Nicotine use disorder is functionally multidimensional, and so is the neural response to smoking-related cues. Here, we disentangled brain responses to two core dimensions associated with smoking-related cues: craving and valence. Using a parametric fMRI design, we investigated the influence of craving- and valence aspects on key addiction brain areas: the amygdala, the ventral striatum, the ACC, the PCC, and the OFC. Our approach enables a more precise mapping of craving-associated brain areas.

Our results show that the chosen brain areas that are robustly recruited by smoking cues in meta-analyses (24-26) were indeed activated by the craving-aspect of the presented cues (Figure 4). Here, it should be noted that smoking cue-reactivity in single studies can be dependent on context (e.g drug availability right after the scanning session) and the subject population (e.g., treatment seeking subjects versus non-treatment seeking subjects) and, therefore, might differ from large-scale meta-analyses (49).

However, when isolating the emotional content of the smoking cues and its influence on neural responses within these ROIs, we observed that amygdala activation is driven by the images’ valence ratings. This valence-driven response in the amygdala was significantly stronger than the craving-driven amygdala response. In contrast, the ACC, ventral striatum, OFC, and PCC, were not (strongly) affected by the emotional content of the presented images. Hence, while ACC, ventral striatum, OFC, and PCC are associated specifically with the urge to smoke, the amygdala responds primarily to the valence dimension of smoking cues, which is a novel finding in the cue-reactivity literature.

This has practical implications for the interpretation of previous studies, the design of future cue-reactivity research, and for the further development of neuronal models of addiction. The amygdala has frequently been reported as activated in smoking cue-reactivity paradigms and has correctly been identified as an important region in addiction (31,32,50). However, previous studies did not disentangle which dimension the amygdala responds to. According to a literature search of the studies included in three cue-reactivity meta-analyses (24-26) only two out of 49 studies (51,52), i.e. only 4% of the studies, accounted for stimulus valence in their study designs. Further, 80% of the included studies used a cue-reactivity block-design with the other studies using an event-related, yet non-parametric design. In neuroimaging, classical block designs are commonly used for because of their greater statistical power compared to, for example, parametric designs (40,41). However, when blocks of craving cues are contrasted with blocks of neutral images, the resulting brain activations reflect differences between these craving cues and neutral images along all functional dimensions of the stimuli and of nicotine dependence. Our results show that when the influence of valence is not accounted for in a cue-reactivity design, amygdala responses will most likely be driven by the valence and not the craving aspect of the presented smoking cues. This can be avoided by, for example, adjusting for valence during stimulus selection or specifically focusing on the craving dimension using a parametric modulator analysis approach. Publicly available smoking cue databases such as SmoCuDa (https://smocuda.github.io/; (46)) provide suitable stimuli for implementing such designs. If this is not possible, the uncertainty regarding which smoking dimension is causing the activations should be kept in mind when interpreting them. These suggestions do not just apply to smoking research, but can also inform other fields that work with fMRI-based cue reactivity paradigms, such as alcohol use disorder (53), heroin use disorder (54), cocaine use disorder (55,56), obesity (57), and gambling disorder (58).

We applied a parametric fMRI design combined with behavioral craving and valence ratings to disentangle core components of craving- and valence-driven brain regions. Such a specific parametric cue-reactivity approach can grant more detailed and precise insights into the complex dynamics of cue-driven craving in the brain, as it allows for focusing on separable stimulus dimension. Using this design, we identified key regions within the addiction network that were more sensitive to and more driven by either the craving or the emotional content of the presented smoking cues. Future studies might also investigate further stimulus dimensions which might be intertwined with craving-related results in key brain areas for addiction, such as salience or arousal. This can help to further specify the functional role of brain regions included in existing neuronal models of addiction (33,59-61). A better functional understanding and more precise mapping of the neural underpinnings of craving might also help to identify more reliable targets for clinical interventions, such as cue-exposure therapy or real-time fMRI neurofeedback. In particular novel brain-based treatment approaches such as neurofeedback rely strongly on the correct selection of target brain areas and signals (62,63). Neurofeedback has been shown to be a promising tool for treating dysfunctional brain signals in substance use disorder (64-66), and in the field of tobacco use disorder, several brain areas have been successfully trained to reduce smoking cue-driven drug craving and to support smoking cessation (67-71).

## Conclusion

Using a parametric cue-reactivity paradigm, we disentangled brain responses to craving and valence dimensions of smoking cues. Our findings suggest that the amygdala responds primarily to the valence component of such cues. This can refine the interpretation of previous reports of amygdala activity during smoking cue-reactivity and help designing future research aimed at teasing apart the many psychological dimensions that comprise nicotine dependence. Also, a more precise mapping of craving-driven brain areas is an important step towards more tailored smoking cessation treatments.

## Statement of Ethics

This study was conducted in accordance with the Declaration of Helsinki and was approved by the local ethics committee (University of Zurich).

## Acknowledgments

The authors thank all volunteers for participating in this study.

## References

1. World Health Organization. WHO report on the global tobacco epidemic. 2017.

2. Samet JM. Tobacco smoking. The leading cause of preventable disease worldwide. Thorac Surg Clin [Internet]. 2013;23(2):103–12. Available from: http://dx.doi.Org/10.1016/j.thorsurg.2013.01.009

3. Lim SS, Vos T, Flaxman AD, Danaei G, Shibuya K, Adair-rohani H, et al. A comparative risk assessment of burden of disease and injury attributable to 67 risk factors and risk factor clusters in 21 regions, 1990 - 201013: a systematic analysis for the Global Burden of Disease Study 2010. 2012;380:1990–2010.

4. Thyrian JR, Panagiotakos DB, Polychronopoulos E, West R, Zatonski W, John U. The relationship between smokers’ motivation to quit and intensity of tobacco control at the population level: A comparison of five European countries. BMC Public Health. 2008;8:1–6.

5. Piasecki TM. Relapse to smoking. Clin Psychol Rev. 2006;26(2):196–215.

6. Piasecki TM, Baker TB. Any further progress in smoking cessation treatment? Nicotine Tob Res. 2001;3(4):311–23.

7. Shiffman S, Brockwell SE, Pillitteri JL, Gitchell JG. Use of Smoking-Cessation Treatments in the United States. Am J Prev Med. 2008;34(2):102–11.

8. Quednow BB, Herdener M. Human pharmacology for addiction medicine: From evidence to clinical recommendations. In: Progress in Brain Research [Internet]. 1st ed. Elsevier B. V.; 2016. p. 227–50. Available from: http://dx.doi.org/10.1016/bs.pbr.2015.07.017

9. Cohen S, Lichtenstein E. Perceived stress, quitting smoking, and smoking relapse. Health Psychol. 1990;9(4):466–78.

10. Doran N, Spring B, McChargue D, Pergadia M, Richmond M. Impulsivity and smoking relapse. Nicotine Tob Res. 2004;6(4):641–7.

11. Gulliver SB, Hughes JR, Solomon U, Dey AN. An investigation of self-efficacy, partner support and daily stresses as predictors of relapse to smoking in self-quitters. Addiction. 1995;90(6):767–72.

12. Killen JD, Fortmann SP. Craving is associated with smoking relapse: Findings from three prospective studies. Exp Clin Psychopharmacol. 1997;5(2):137–42.

13. Bagot KS, Heishman SJ, Moolchan ET. Tobacco craving predicts lapse to smoking among adolescent smokers in cessation treatment. Nicotine Tob Res. 2007;9(6):647–52.

14. Elwafi HM, Witkiewitz K, Mallik S, Iv TAT, Brewer JA. Mindfulness training for smoking cessation: Moderation of the relationship between craving and cigarette use. Drug Alcohol Depend [Internet]. 2013;130(1-3):222–9. Available from: http://dx.doi.Org/10.1016/j.drugalcdep.2012.11.015

15. Russell MA. Nicotine replacement: the role of blood nicotine levels, their rate of change, and nicotine tolerance. Prog Clin Biol Res. 1988;261:63–94.

16. Sayette MA, Shiffman S, Tiffany ST, Niaura RS, Martin CS, Schadel WG. The measurement of drug craving. Addiction. 2000;95(8):189–210.

17. Gilbert HM, Warburton DM. Craving: A problematic concept in smoking research. Addict Res Theory. 2000;8(4):381–97.

18. Shiffman S, Engberg JB, Paty JA, Perz WG, Gnys M, Kassel JD, et al. A day at a time: Predicting smoking lapse from daily urge. J Abnorm Psychol. 1997;106(1):104–16.

19. Hughes JR. Tobacco withdrawal in self-quitters. J Consult Clin Psychol. 1992;60(5):689–97.

20. Tiffany ST, Drobes DJ. The development and initial validation of a questionnaire on smoking urges. Br J Addict. 1991;86(ll):1467–76.

21. Ferguson SG, Shiffman S. The relevance and treatment of cue-induced cravings in tobacco dependence. J Subst Abuse Treat [Internet]. 2009;36(3):235–43. Available from: http://dx.doi.org/10.1016/jjsat.2008.06.005

22. Carter BL, Tiffany ST. Meta-analysis of cue-reactivity in addiction research. Addiction. 1999;94(3):327^40.

23. Niaura R, Shadel WG, Abrams DB, Monti PM, Rohsenow DJ, Sirota A. Individual differences in cue reactivity among smokers trying to quit: Effects of gender and cue type. Addict Behav. 1998;23(2):209–24.

24. Chase HW, Eickhoff SB, Laird AR, Hogarth L. The neural basis of drug stimulus processing and craving: An activation likelihood estimation meta-analysis. Biol Psychiatry [Internet]. 2011;70(8):785–93. Available from: http://dx.doi.Org/10.1016/j.biopsych.2011.05.025

25. Kuhn S, Gallinat J. Common biology of craving across legal and illegal drugs - a quantitative meta-analysis of cue-reactivity brain response. Eur J Neurosci. 2011;33(7):1318–26.

26. Tang DW, Fellows LK, Small DM, Dagher A. Food and drug cues activate similar brain regions: A meta-analysis of functional MRI studies. Physiol Behav [Internet]. 2012;106(3):317–24. Available from: http://dx.doi.Org/10.1016/j.physbeh.2012.03.009

27. Koob GF, Volkow ND. Neurobiology of addiction O: a neurocircuitry analysis. The Lancet Psychiatry [Internet]. 2016;3(8):760–73. Available from: http://dx.doi.org/10.1016/S2215-0366(16)00104-8

28. Schultz W, Apicella P, Scarnati E, Ljungberg T. Neuronal activity in monkey ventral striatum related to the expectation of reward. J Neurosci. 1992;12(12):4595–610.

29. Pagnoni G, Zink CF, Montague PR, Berns GS. Activity in human ventral striatum locked to errors of reward prediction. Nat Neurosci. 2002;5(2):97–8.

30. Gallagher M, Chiba AA. The amygdala and emotion. Curr Opin Neurobiol. 1996;6(2):221–7.

31. Everitt BJ, Parkinson JA, Olmstead MC, Arroyo M, Robledo P, Robbins TW. Associative processes in addiction and reward. The role of amygdala-ventral striatal subsystems. Ann N Y Acad Sci. 1999;877:412–38.

32. Ronald E. See, Rita A. Fuchs, Christpoher C. Ledford, Joselyn McLaughlin. Drug Addiction, Relapse, and the Amygdala. Ann N Y Acad Sci. 2003;985:294–307.

33. Kalivas PW, Volkow ND. The neural basis of addiction: A pathology of motivation and choice. Am J Psychiatry. 2005;162(8):1403–13.

34. Schoenbaum G, Roesch MR, Stalnaker TA. Orbitofrontal cortex, decision-making and drug addiction. Trends Neurosci. 2006;29(2):116–24.

35. Brody AL, Mandelkern MA, Olmstead RE, Jou J, Tiongson E, Allen V, et al. Neural Substrates of Resisting Craving During Cigarette Cue Exposure. Biol Psychiatry. 2007;62(6):642–51.

36. Janes AC, Pizzagalli DA, Richardt S, Frederick B de B, Chuzi S, Pachas G, et al. Brain Reactivity to Smoking Cues Prior to Smoking Cessation Predicts Ability to Maintain Tobacco Abstinence. Biol Psychiatry [Internet]. 2010;67(8):722–9. Available from: http://dx.doi.Org/10.1016/j.biopsych.2009.12.034

37. Allenby C, Falcone M, Wileyto EP, Cao W, Bernardo L, Ashare RL, et al. Neural cue reactivity during acute abstinence predicts short-term smoking relapse. Addict Biol. 2020;25(2):1–9.

38. Phan KL, Wager T, Taylor SF, Liberzon I. Functional neuroanatomy of emotion: A meta-analysis of emotion activation studies in PET and fMRI. Neuroimage. 2002;16(2):331–48.

39. Costafreda SG, Brammer MJ, David AS, Fu CHY. Predictors of amygdala activation during the processing of emotional stimuli: A meta-analysis of 385 PET and fMRI studies. Brain Res Rev. 2008;58(1):57–70.

40. Liu TT, Frank LR, Wong EC, Buxton RB. Detection power, estimation efficiency, and predictability in event-related fMRI. Neuroimage. 2001;13(4):759–73.

41. Miezin FM, Maccotta L, Ollinger JM, Petersen SE, Buckner RL. Characterizing the hemodynamic response: effects of presentation rate, sampling procedure, and the possibility of ordering brain activity based on relative timing. Neuroimage. 2000;11(6 Pt 1):735–59.

42. American Psychiatric Association. Diagnostic and Statiatical Manual of Mental Disorders, Fifth Edition. 2013.

43. Quednow BB, Kühn KU, Hoenig K, Maier W, Wagner M. Prepulse inhibition and habituation of acoustic startle response in male MDMA (‘Ecstasy’) users, cannabis users, and healthy controls. Neuropsychopharmacology. 2004;29(5):982–90.

44. Todd F. Heatherton, Lynn T. Kozlowski, Richard C. Frecker, Karl-Olov Fagerstrom. The Fagerstrom Test for Nicotine Dependence: a revision of the Fagerstrom Tolerance Questionnaire. Br J Addict [Internet]. 1991;86:1119–27. Available from: https://ai2-s2-pdfs.s3.amazonaws.com/74c8/dd44c488807e054a5ed8711f6bc7b2fbeaea.pdf

45. Sanderson Cox, Stephen T. Tiffany, L. Evaluation of the brief questionnaire of smoking urges (QSU-brief) in laboratory and clinical settings. Nicotine Tob Res [Internet]. 2001;3(1):7–16. Available from: https://academic.oup.com/ntr/article-lookup/doi/10.1080/14622200124218

46. Manoliu A, Haugg A, Sladky R, Hulka L, Kirschner M, Bruhl AB, et al. SmoCuDa: A Validated Smoking Cue Database to Reliably Induce Craving in Tobacco Use Disorder. Eur Addict Res. 2020; (in press)

47. Lang PJ, Bradley MM, Cuthbert BN. International Affective Picture System (IAPS): Technical Manual and Affective Ratings. NIMH Cent Study Emot Atten. 1997;1:39–58.

48. Gilbert DG, Rabinovich NE. International Smoking Image Series, Version1.2.1999.

49. Wilson SJ, Sayette MA, Fiez JA. Prefrontal responses to drug cues: A neurocognitive analysis. Nat Neurosci. 2004;7(3):211–4.

50. Roberts AJ, Koob GF. The Neurobiology of Addiction. Alcohol Heal Res World. 1997;21(2):101–6.

51. Filbey FM, Claus E, Audette AR, Niculescu M, Banich MT, Tanabe J, et al. Exposure to the taste of alcohol elicits activation of the mesocorticolimbic neurocircuitry. Neuropsychopharmacology. 2008;33(6):1391–401.

52. Franklin T, Wang Z, Suh JJ, Hazan R, Cruz J, Li Y, et al. Effects of varenicline on smoking cue-triggered neural and craving responses. Arch Gen Psychiatry. 2011;68(5):516–26.

53. Schacht JP, Anton RF, Myrick H. Functional neuroimaging studies of alcohol cue reactivity: A quantitative meta-analysis and systematic review. Addict Biol. 2013;18(1):121–33.

54. Yang Z, Xie J, Shao YC, Xie CM, Fu LP, Li DJ, et al. Dynamic neural responses to cue-reactivity paradigms in heroin-dependent users: An fMRI study. Hum Brain Mapp. 2009;30(3):766–75.

55. Wilcox CE, Teshiba TM, Merideth F, Ling J, Mayer AR. Enhanced cue reactivity and fronto- striatal functional connectivity in cocaine use disorders _. Drug Alcohol Depend [Internet]. 2011;115(1-2):137–44. Available from: http://dx.doi.Org/10.1016/j.drugalcdep.2011.01.009

56. Goldstein RZ, Alia-klein N, Tomasi D, Honorio J, Maloney T, Woicik PA, et al. Anterior cingulate cortex hypoactivations to an emotionally salient task in cocaine addiction. 2009;2.

57. Boswell RG, Kober H. Food cue reactivity and craving predict eating and weight gain: A meta-analytic review. Obes Rev. 2016;17(2):159–77.

58. Limbrick-Oldfield EH, Mick I, Cocks RE, McGonigle J, Sharman SP, Goldstone AP, et al. Neural substrates of cue reactivity and craving in gambling disorder. Transl Psychiatry. 2017;7(1):1–10.

59. Robinson TE, Berridge KC. The neural basis of drug craving: An incentive-sensitization theory of addiction. Brain Res Rev. 1993;18(3):247–91.

60. Berridge KC, Robinson TE. Liking, Wanting and the Incentive-Sensitization Theory. Am Psychol. 2016;71(8):670–9.

61. Volkow ND, Koob GF, McLellan AT. Neurobiologic advances from the brain disease model of addiction. N Engl J Med. 2016;374(4):363–71.

62. Sitaram R, Ros T, Stoeckel LE, Haller S, Scharnowski F, Lewis-Peacock J, et al. Closed-loop brain training: the science of neurofeedback. Nat Neurosci [Internet]. 2016; Available from: http://dx.doi.org/10.1038/nrn.2016.164

63. Weiskopf N. Real-time fMRI and its application to neurofeedback. Neuroimage [Internet]. 2012;62(2):682–92. Available from: http://dx.doi.Org/10.1016/j.neuroimage.2011.10.009

64. Kirschner M, Sladky R, Haugg A, Stämp P, Jehli E, Hodel M, et al. EBioMedicine Self-regulation of the dopaminergic reward circuit in cocaine users with mental imagery and neurofeedback. 2018;37:489–98.

65. Karch S, Keeser D, Hümmer S, Paolini M, Kirsch V, Karali T, et al. Modulation of craving related brain responses using real-time fMRI in patients with alcohol use disorder. PLoS One. 2015;10(7).

66. Weiss F, Aslan A, Zhang J, Gerchen MF, Gerchen MF, Gerchen MF, et al. Using mind control to modify cue-reactivity in AUD: The impact of mindfulness-based relapse prevention on real-time fMRI neurofeedback to modify cue-reactivity in alcohol use disorder: A randomized controlled trial. BMC Psychiatry. 2020;20(1):1–11.

67. Hartwell KJ, Hanlon CA, Li X, Borckardt JJ, Canterberry M, Prisciandaro JJ, et al. Individualized real-time fMRI neurofeedback to attenuate craving in nicotine-dependent smokers. J Psychiatry Neurosci. 2016;41(1):48–55.

68. Hanlon CA, Hartwell KJ, Canterberry M, Li X, Owens M, LeMatty T, et al. Reduction of cue-induced craving through realtime neurofeedback in nicotine users: The role of region of interest selection and multiple visits. Psychiatry Res - Neuroimaging [Internet]. 2013;213(1):79–81. Available from: http://dx.doi.Org/10.1016/j.pscychresns.2013.03.003

69. Canterberry M, Hanlon CA, Hartwell KJ, Li X, Owens M, LeMatty T, et al. Sustained reduction of nicotine craving with real-time neurofeedback: Exploring the role of severity of dependence. Nicotine Tob Res. 2013;15(12):2120–4.

70. Kim D-Y, Yoo S-S, Tegethoff M, Meinlschmidt G, Lee J-H. The Inclusion of Functional Connectivity Information into fMRI-based Neurofeedback Improves Its efficacy in the Reduction of Cigarette Cravings. J Cogn Neurosci. 2015;27(8):1552–72.

71. Karch S, Paolini M, Gschwendtner S, Jeanty H, Reckenfelderbäumer A, Yaseen O, et al. Real-time fMRI neurofeedback in patients with tobacco use disorder during smoking cessation: Functional differences and implications of the first training session in regard to future abstinence or relapse. Front Hum Neurosci. 2019;13(March):1–17.

